# Use, reuse or discard: quantitatively defined variance in N95 respirator integrity following vaporized hydrogen peroxide decontamination during the COVID-19 pandemic

**DOI:** 10.1101/2020.08.18.20177071

**Authors:** Carly Levine, Courtney Grady, Thomas Block, Harry Hurley, Riccardo Russo, Blas Peixoto, Alexis Frees, Alejandro Ruiz, David Alland

## Abstract

**Background:** COVID-19 has stretched the ability of many institutions to supply needed personal protective equipment, especially N95 respirators. N95 decontamination and reuse programs provide one potential solution to this problem. Unfortunately, a comprehensive evaluation of the effects of decontamination on the integrity of various N95 models using a quantitative fit test (QTFT) approach is lacking.

**Aims:** 1) To investigate the effects of up to eight rounds of vaporized H_2_O_2_ (VHP) decontamination on the integrity of N95 respirators currently in use in a hospital setting. 2) To examine if N95 respirators worn by one user can adapt to the face shape of a second user with no compromise of integrity following VHP decontamination.

**Methods:** The PortaCount Pro+ Respirator Fit Tester Model 8038 was used to quantitatively define the integrity, measured by fit, of N95 respirators following decontamination with VHP.

**Findings:** There was an observable downward trend in the integrity of Halyard Fluidshield 46727 N95 respirators throughout eight cycles of decontamination with VHP. The integrity of 3M 1870 N95 respirators was significantly reduced after the respirator was worn, decontaminated with VHP, and then quantitatively fit tested on a second user. Furthermore, we uncovered inconsistencies between qualitative fit test and QTFT results that may have strong implications on the fit testing method used by institutions.

**Conclusions:** Our data revealed variability in the integrity of different N95 models after VHP decontamination and exposed potential limitations of N95 decontamination and reuse programs.

## INTRODUCTION

As of August 2020, over 21.7 million people have been infected and more than 770,000 have died from COVID-19 worldwide [1]. Healthcare workers are at high risk of contracting COVID-19 [2–4], making personal protective equipment (PPE), including N95 respirators, critical for their protection. Many hospitals have universally implemented the use of N95 respirators during routine care to limit exposure to mild and asymptomatic individuals and during aerosol-producing procedures, such as intubation and mechanical ventilation [5,6]. This increase in use has left many hospitals struggling to maintain adequate stock of N95 respirators in the face of increasing supply chain shortages [7–10].

Decontamination and reuse of N95 respirators is a potential solution for alleviating respirator scarcity during the COVID-19 pandemic as per the Occupational Safety and Health Administration’s (OSHA) recently updated guidelines [11]. Previous studies have validated the use of vaporized hydrogen peroxide (VHP) [12], moist heat [13], dry heat [14], or ultraviolet germicidal radiation [15] for decontamination of N95 respirators. The integrity of decontaminated N95 respirators, measured by fit, must be maintained following processing. Proper fit is defined as an intact, airtight seal against the user’s face that can be measured using either a qualitative fit test (QLFT) or a quantitative fit test (QTFT), as defined by Appendix A of the OSHA Standard 1920.134 [16]. A QTFT more accurately identifies proper fit than a QLFT [17,18]. However, an N95 respirator that is tested using a QTFT cannot be subsequently worn for future protection because this procedure requires puncturing a hole into the respirator to assess fit. On the other hand, N95 respirators examined via a QLFT can be kept and used by the wearer after the test is completed.

Current decontamination studies only use a manikin head form to quantitatively assess fit factor and fail to evaluate all respirator models used in hospital settings [19,20]. However, product scarcity has required the use of many different N95 models, most of which have not been evaluated after decontamination. Additionally, it can be challenging to develop and maintain systems which ensure that respirators are returned to their original user, as opposed to returning the same model but from a different user post-decontamination. Yet, the integrity of N95 respirators worn by multiple persons has not been investigated.

Here, we first evaluated the number and type of N95 respirators qualitatively fit tested and distributed by University Hospital (UH) in Newark, NJ before and during the COVID-19 pandemic. We then used QTFTs to assess the integrity of 4/7 of these N95 models following sequential rounds of decontamination with VHP. The integrity of decontaminated N95 respirators was then tested on a second user using a QTFT approach. Finally, we evaluated the reliability of qualitative fit testing on models that we found were hard to fit quantitatively by comparing QLFT vs. QTFT results. Our findings revealed that the majority of N95 respirators evaluated were able to withstand the VHP decontamination process and that these N95 respirators could be returned to new users with no significant decrease in integrity. However, we did observe exceptions to these findings that may have strong implications on which respirator models are suitable for decontamination and reuse programs. Furthermore, differences in QLFT and QTFT results for certain respirator models uncovered potential weaknesses in using QLFTs for measuring the protective ability of N95 respirators.

## METHODS

### Human subjects approval

Experiments involving fit testing and decontamination of N95 respirators was part of the public health response to the COVID-19 pandemic and were thus considered exempt from institutional review board approval. All participants gave informed consent prior to participating in any experiments.

### N95 Decontamination

Respirators were decontaminated using VHP which was delivered via the Steris VHP system (Steris Life Sciences, Mentor, OH). Respirator hanging and decontamination was similar to previously published studies [12].

### Fit Testing

Fit testing was administered following OSHA standard Appendix A to 1920.134 with minor modifications: 1) N95 respirators tested using both QLFTs and QTFTs underwent QLFTs first, 2) QLFT users were blinded to the order by which qualitative testing was administered (sweet followed by bitter). All users were deemed medically able to complete fit testing beforehand.

### Qualitative fit testing

The 3M Qualitative Fit Test Apparatus FT-30, Bitter (denatonium benzoate) 1 kit and 3M Qualitative Fit Test Apparatus FT-10, Sweet (saccharine) kit (3M, Saint Paul, MN) were used for QLFTs. These kits included a hood, collar, and nebulizers which were used to aerosolize the tasting solution. All users were sensitive to tasting agents.

### Quantitative fit testing

The QTFTs were administered using a PortaCount Pro+ Respirator Fit Tester Model 8038 with an N95-Companion™ Fit N95 Tester Model 8026 Particle Generator (TSI, Shoreview, MN). Respirators were punctured with custom grommet sampling probes to connect a sampling tube between the inside of an N95 respirator and the PortaCount Pro+ machine. Sodium chloride tablets were used for particle generation. All testing was conducted above the minimum recommendations of 70 ambient particles/cc and a passing test required a fit factor of > 100. Fit factors were calculated by the apparatus from the ratio of particles outside to particles inside the respirator.

### Second user N95 respirators

Respirators that had previously failed a QLFT were termed “lightly used” because even though the nosepiece had been fit and shaped to a user’s face, the respirator did not undergo the extended wear that is representative of a long hospital shift. These lightly used N95 respirators were used for second user experiments.

### Respirator sizing

All respirators used except Kimberly Clark/Halyard Fluidshield (Halyard Fluidshield) and 3M 1860 were available in one size only. Halyard Fluidshield N95 respirators were available in small (46827) and regular (46727) sizes. 3M 1860 N95 respirators were also available in small (1860S) and regular (1860) sizes. For these models, the appropriate size was determined for each participant using a QTFT before the start of the study. The correct size was then used for each subject in all experiments.

### Statistical analysis

To analyze the effect of decontamination on N95 integrity we used a one tailed Kruskal-Wallis test. A one-tailed Mann-Whitney U test was used to assess the differences between N95 respirators worn by one user and N95 respirators worn by a second user following decontamination. All statistical analysis was run using Prism 8 software.

## RESULTS

### Respirator models and fit testing have increased during the COVID-19 pandemic in a hospital setting

To define the scope of our targeted decontamination study, we examined the use of N95 respirators at UH before and during the COVID-19 pandemic. The frequency of QLFTs in 2020 from January to May was about tenfold greater than the average monthly frequency from 2014 to 2019 (Figure 1A). Over 99% of fit testing conducted before 2020 was on one of four N95 models, Halyard Fluidshield 46727/46827 (59.59%), 3M 1860/3M1860S (25.75%), 3M 1870 (8.68%), and Cardinal Health (5.79%) (Figure 1B). In addition to these models, Gerson 2130 (15.4%), Gerson 1730 (5.38%), and 3M 9210 (4.93%) were introduced during January to May 2020 (Figure 1B). However, Halyard Fluidshield 46727/46827 (21.97%), 3M 1860/3M 1860S (17.64%), 3M 1870 (13%), and Cardinal Health (21.23%) still comprised the majority of the respirators distributed (Figure 1B). Overall, we observed an increase in the diversity, fit test frequency, and use of N95 respirators during the COVID-19 pandemic.

**Figure 1:**
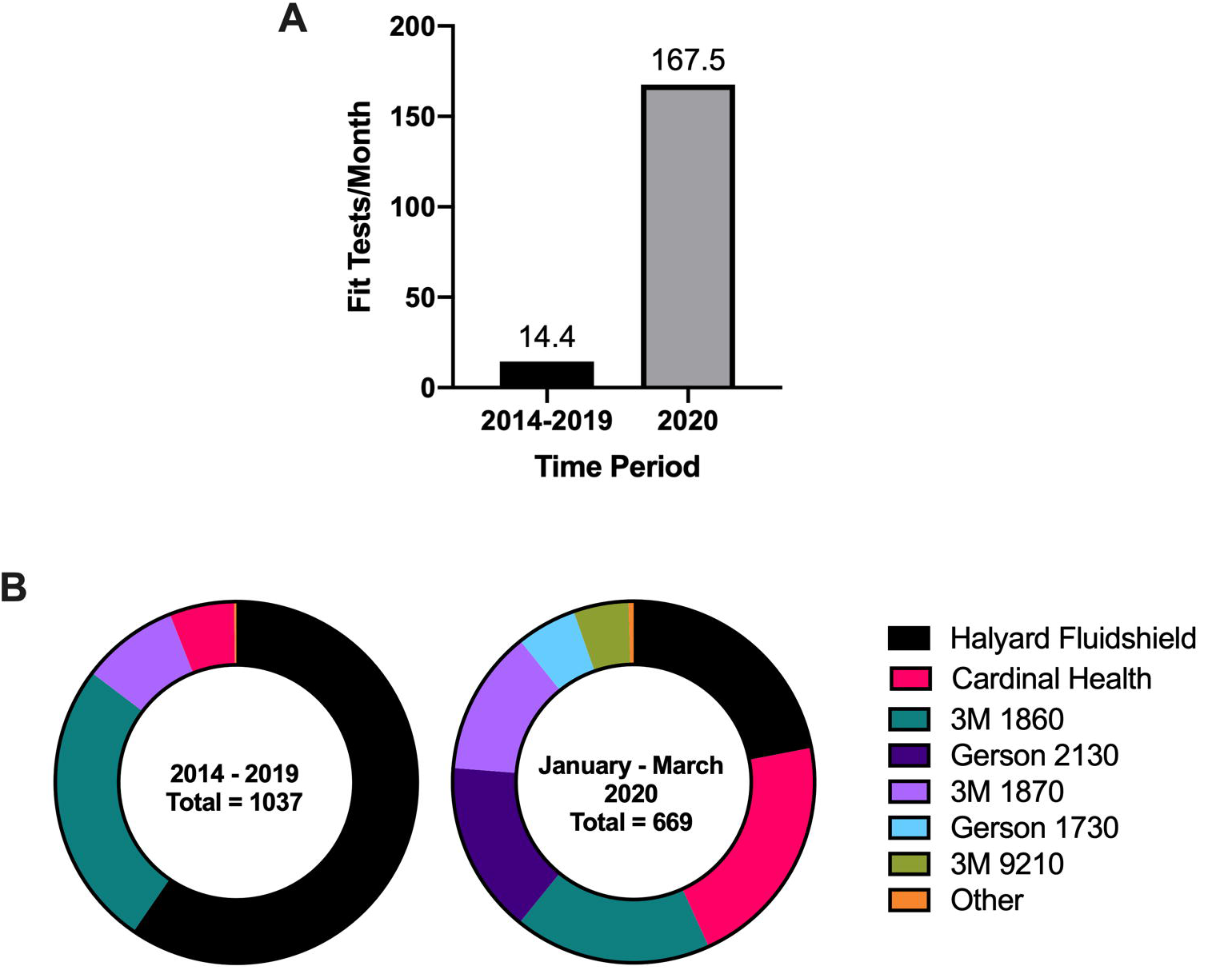
Fit test frequency and distribution of N95 respirators by UH. **A)** The frequency of QLFTs increased tenfold and the **B)** diversity of N95 models expanded since the beginning of the COVID-19 pandemic.

### Variation in quantitative fit testing across different N95 models

Prior to any VHP decontamination, we conducted QTFTs on the seven N95 models used at UH during the COVID-19 pandemic (Figure 2). We defined a QTFT fit factor of ≥ 100 to be a passing value. Users who passed a QTFT were considered certified to use the N95 model tested. Respirator models with high frequency of passing across different users were 3M 1860/3M 1860S, 3M 1870, 3M 9210, and Halyard Fluidshield 46727, which had passing rates of 71%, 100%, 75%, and 80%, respectively. The other models tested had much lower QTFT passing rates across different users with Cardinal Health, Gerson 1730, and Gerson 2130 having passing rates of 0%, 10%, and 11%, respectively. These models may work well with face types and sizes not representative in our volunteer population. However, for our studies we defined these last three models as “hard-to-fit” and did not include them in the decontamination experiments.

**Figure 2:**
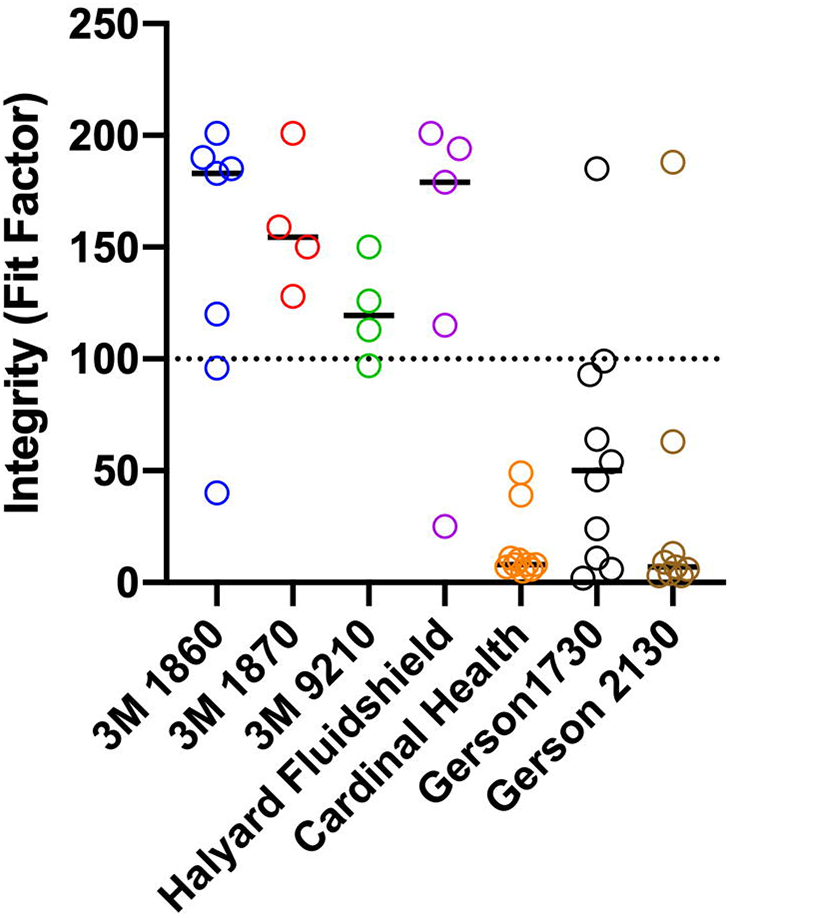
Quantitative fit testing of N95 respirators commonly used at UH between January and May 2020. These data defined quantitatively hard-to-fit N95 models: Cardinal Health, Gerson 1730, and Gerson 2130. A fit factor > 100 was considered a pass (dotted line). Bars represent the median. 3M 1860/3M 1806S n = 7, 3M 1870 n = 4, 3M 9210 n = 4, Halyard Fluidshield 46727 n = 5, Cardinal Health n = 10, Gerson 1730 n = 10, Gerson 2130 n = 9.

### Decontamination with VHP does not affect the integrity of 3M 1860/3M 1860S, 3M 1870, 3M 9210, but may reduce the integrity of Halyard Fluidshield 46727

To evaluate the effect of VHP decontamination on N95 respirator integrity, new unworn N95 respirators were decontaminated consecutively for up to eight cycles. Following each decontamination cycle, a subset was removed for quantitative fit testing. Availability and supply limitations of N95 respirators influenced which models were evaluated at each decontamination cycle and only cycles with at least n = 3 were analyzed. Both 3M 1860/3M 1860S and 3M 1870 N95 respirators maintained integrity following up to eight and six cycles of VHP decontamination, respectively (Figure 3A and 3B). There was also no significant difference in the integrity of 3M 9210 following up to seven decontamination cycles (Figure 3C). We observed a clear downward trend in the integrity of Halyard Fluidshield 46727 N95 respirators throughout eight decontamination cycles (Figure 3D). However, due to the limited number of respirators during this critical time we were unable to detect any significant differences in the data. Importantly, we did not notice any defects in the elastic headbands, nor did we observe any corrosion on the metal nosepiece and staples following eight cycles with VHP which has also been validated by others [19]. Our observations suggest that some but not all N95 models are appropriate to include in respirator decontamination and reuse programs and warrant further study.

**Figure 3:**
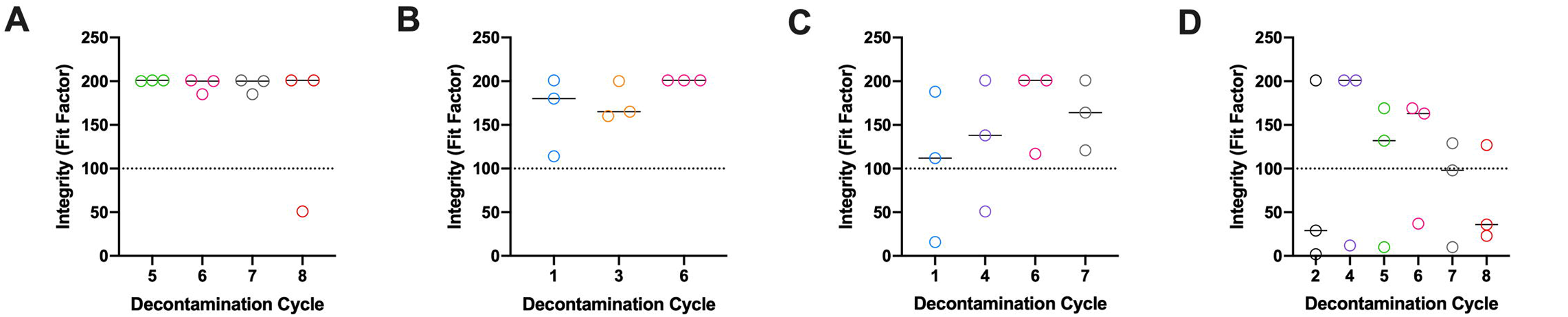
Decontamination with VHP does not affect the integrity of A) 3M 1860, B) 3M 1870, C) 3M 9210, but potentially reduces the integrity of D) Halyard Fluidshield 46727. No significant differences were observed between decontamination cycles for any model when analyzed using a one tailed Kruskal-Wallis test. However, there was an observable downward trend in the integrity of Halyard Fluidshield 46727 throughout eight cycles of decontamination. A passing value was set as a fit factor ≥ 100 (dotted line). Only decontamination cycles with at least n = 3 were considered for analysis. Bars represent the median.

### Decreased integrity of 3M 1870 N95 respirators is observed when the respirator is lightly worn and then fit tested by a second user

Not all N95 models will be compatible with all face types and sizes. Respirators that fail QLFTs are generally discarded, but that does not mean they will not provide protection to a different user; it means that the respirator and face type are incompatible. Instead of letting these respirators go to waste, we decontaminated them with VHP and then investigated the flexibility of their face-sealing capacity by assessing integrity on a second user. These second users had previously passed a QTFT on the model (and size where appropriate) being tested. There was a significant decrease in the integrity of 3M 1870 N95 respirators when the respirator was lightly worn, decontaminated for six cycles with VHP, and fit tested by a second user compared to a respirator of the same model and size that had undergone the same number of decontamination cycles but was never previously used (Figure 4B). No significant differences were observed in the integrity of 3M 1860/3M 1860S, 3M 9210, and Halyard Fluidshield 46727 N95 respirators when fit tested by a second user following multiple rounds of decontamination (Figure 4 A, C, D). Again, due to the limited supply of N95 respirators we were unable to assess the differences in integrity between first user and second user for all cycles and models (Figure S1 A-D).

**Figure 4:**
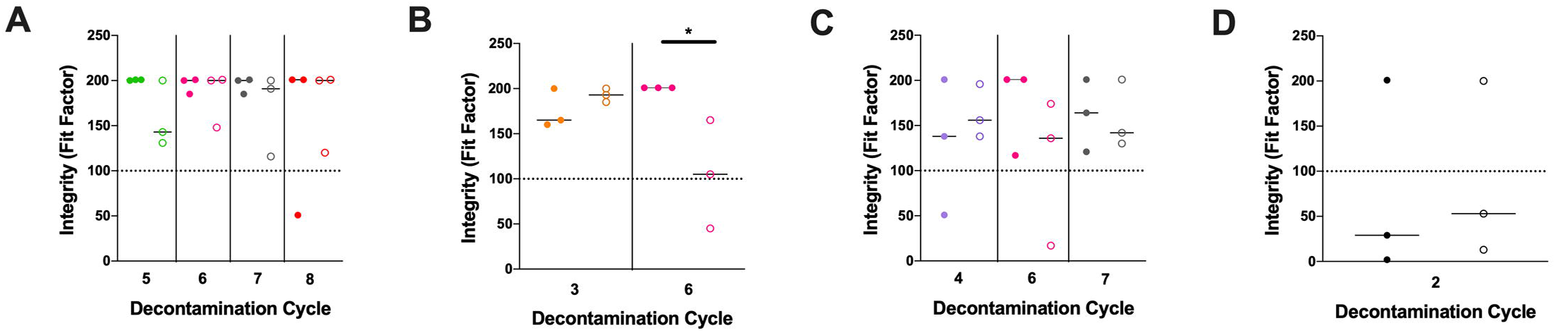
Comparison of the integrity of N95 respirators that were not previously worn to N95 respirators that were lightly worn. Fit test data from our earlier decontamination studies on new N95 respirators (closed circles) were used to evaluate the effect of light wear (open circles) on N95 respirator integrity following VHP decontamination. **A)** A nonsignificant downward trend in the integrity of 3M 1860/3M 1860S N95 respirators is observed following five, six, and seven decontamination cycles when the N95 respirator was previously worn. **B)** 3M 1870s that were lightly worn, decontaminated for six cycles, and fit on a new second user had a significant decrease in integrity compared to 3M 1870 N95 respirators that were decontaminated for six cycles but not previously worn. No significant trends were found in the integrity of **C)** 3M 9210 and **D)** Halyard Fluidshield 46727 when the N95 respirator was previously worn. Only decontamination cycles with at least n = 3 were considered for analysis. A passing value was set as a fit factor ≥ 100 (dotted line). Bars represent the median. A one tailed Mann-Whitney U test was used for statistical analysis (*, p = 0.025).

### Discrepancies between qualitative and quantitative fit testing are apparent when evaluating Gerson 1730

We expanded our study to examine the possibility of inconsistencies between QLFT and QTFT results from hard-to-fit N95 models. Using both sweet and bitter QLFT measurements followed by a QTFT, we determined that 5/6 participants who passed at least one qualitative fit test were unable to pass quantitatively when testing the Gerson 1730 N95 respirator (Table I). Unfortunately, we were unable to find enough participants able to qualitatively fit either Gerson 2130 or Cardinal Health N95 respirators and therefore they were not included in this study (Table SI). The inconsistency between qualitative and quantitative fit testing results calls to question the reliability of QLFTs for measuring the capacity of N95 respirators to protect against aerosol exposure.

**Table I:**
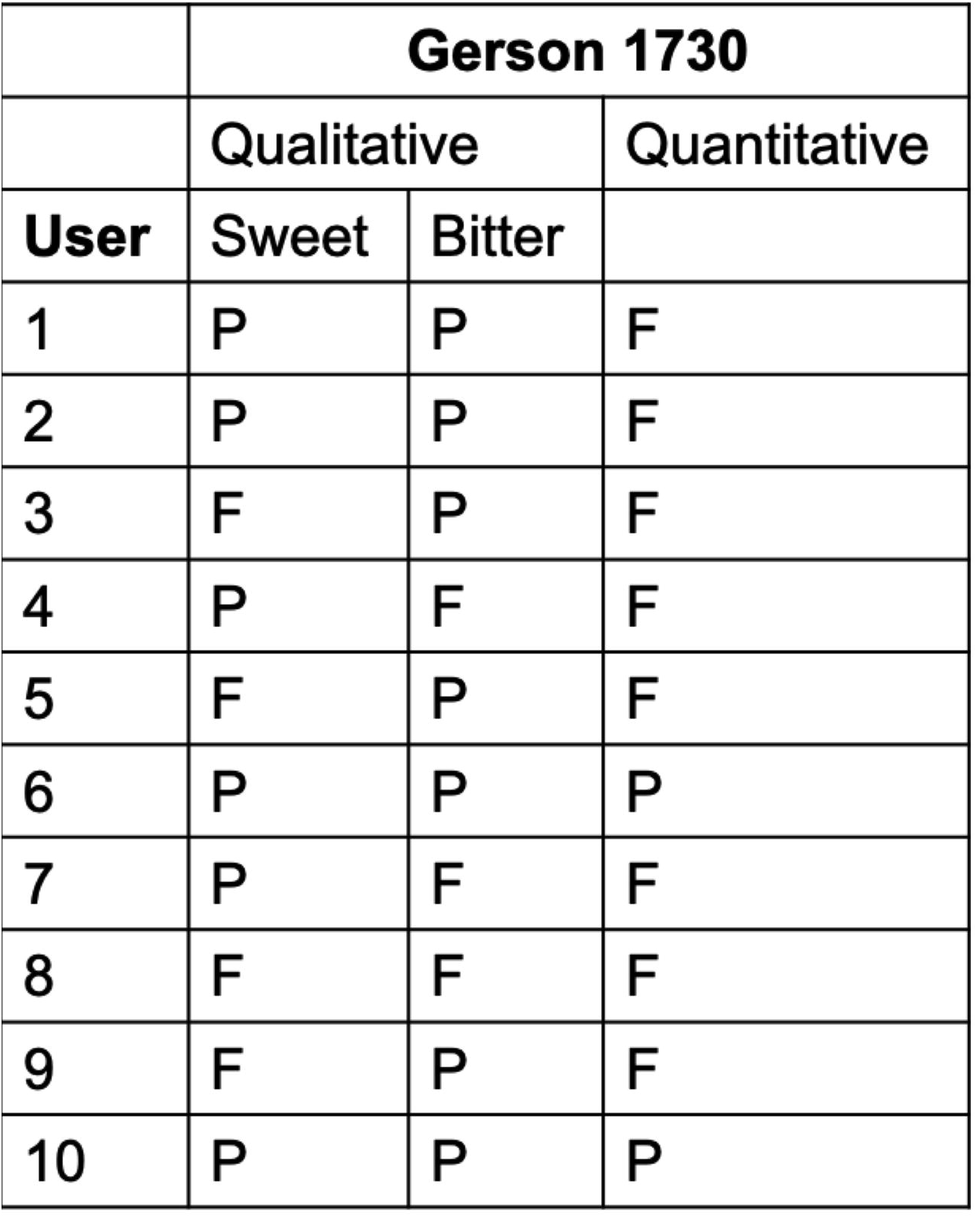
Frequent inconsistencies between QLFT and QTFT results were uncovered when fit tested on Gerson 1730 N95 respirators. P = pass, F = fail

## DISCUSSION

COVID-19 was officially declared a pandemic on 11^th^ March 2020 and has since resulted in an overburdened healthcare system that is struggling to maintain adequate PPE, especially N95 respirators. Many clinical settings have implemented decontamination and reuse programs in response to these shortages. However, recent evaluations of N95 integrity following decontamination have only assessed a single N95 model, 3M 1860/3M 1860S [12–15], and therefore do not account for the total variety of N95 respirators currently in use. This lack of validation means health care workers could be at an increased risk of receiving inadequate PPE. Thus, in this study we quantitatively evaluated the integrity of multiple N95 models currently in use at UH following decontamination with VHP.

From January to May 2020, UH increased qualitative fit testing over tenfold and expanded the models of N95 respirators distributed compared to six years prior. Using unworn N95 respirators, we examined the integrity of these models after decontamination with VHP and found that 3M 1860/3M 1860S, 3M 1870, and 3M 9210 did not exhibit any noticeable decrease of integrity. Although not significant, we did observe a downward trend in the integrity of Halyard Fluidshield 46727 N95 respirators over the course of five decontamination cycles, highlighting the importance of further studies.

It is crucial that decontamination and reuse programs are able to rapidly turnaround clean N95 respirators. One potential time saving approach is to return N95 respirators to new users instead of having to sort and return respirators to their initial user. To assess the adaptability of N95 respirators to a new face we quantitatively fit tested lightly worn respirators on a second user following decontamination with VHP. Our results were limited to respirators that were initially only worn for a QLFT and are thus not representative of the wear and tear associated during an extended hospital shift. Despite these limitations, the QTFT values were lower for 3M 1870 N95 respirators when they were lightly worn and fit tested by a second user compared to the other respirator models tested. It is also possible that some respirators may be less tolerant for reuse even by the same person, however we were unable to assess that in this study. Although, others have found a significant association between number of shifts N95 respirators were worn by a single user and failed QLFTs [21]. These data bring to light a significant obstacle for decontamination and reuse programs.

A major unexpected result in this study was the inability to find many users who passed QTFTs on Gerson 1730, Gerson 2130, and Cardinal Health N95 models, despite these respirators representing 42% of passing QLFTs at UH. We therefore wondered if differences existed between QTFT and QLFT results specifically on these hard-to-fit models. Of the ten participants we recruited for QLFTs, six were able to pass at least one of the tasting challenges when qualitatively fit tested on Gerson 1730. Interestingly, only 1/6 of these participants was able to pass on the Gerson 1730 using a QTFT. This discrepancy between QLFTs and QTFTs may be attributed to taste insensitivity which has been shown to increase false positive fit testing [18,22] and could also be a symptom of COVID-19 infection [23–25]. Furthermore, anecdotal reports have indicated issues with competing taste profiles such as previously eaten foods or disinfectants used to clean the hood that may further complicate QLFT results. Together these observations suggest the administration of QTFTs may be warranted for fit testing these hard-to-fit models and should be used to assess inconsistencies between QLFT on other N95 models.

We acknowledge that this study has several limitations. All fit testing methods can have false pass rates of up to 11% [17]. The number of N95 respirators fit tested after each decontamination cycle was restricted by supply availability and resulted in unequal sample sizes between groups. Finally, when assessing the integrity of N95 respirators fit on a second user, the original N95 respirator was not worn during a long shift and therefore may not be representative of all respirators included in decontamination and reuse programs.

## CONCLUSIONS

Decontamination and reuse of 3M 1860/3M 1860S, 3M 1870, and 3M 9210 N95 respirators is a potential solution to N95 respirator supply shortages. Further studies must address the downward trends observed in the integrity of Halyard Fluidshield 46727 N95 respirators after decontamination with VHP. Caution should be taken when returning 3M 1870 to a second user following VHP decontamination. Finally, the lack of consistency between QLFT and QTFT results may have far reaching consequences on the type of fit test administered by institutions when determining which respirator is best for protection against aerosolized pathogens.

## Data Availability

All data is available within the manuscript and supplementary material.

## ACKNOWLEDGEMENTS

We would like to thank Dr. Jessica McCormick Ell (National Institutes of Health), Dr. Anthony Gresko (Rutgers University), Guillaume Delmas (Rutgers University), Brian Eggert (Rutgers University), Dr. Mark Einstein (UH), Dr. Debra Chew (Rutgers NJMS), and all of the other members of The Center for COVID-19 Response and Pandemic Preparedness team who helped get the N95 decontamination and reuse program up and running. We would like to thank Safia Amatullah (UH), Lee Clark (UH), and Jo Ellen Harris (UH) for coordinating N95 respirator delivery to the decontamination site. We would also like to thank Dr. Pradeep Kumar (Rutgers NJMS), Dr. Padmapriya Banada (Rutgers NJMS), Dr. Sukalyani Banik (Rutgers NJMS), Heta Parmar (Rutgers NJMS), Skarleth Moran (Rutgers NJMS), Shraddha Suryavanshi (Rutgers NJMS), and Kaheerman Saibire (Rutgers NJMS) for volunteering to participate in our fit testing studies.

## CONFLICT OF INTEREST STATEMENT

None declared.

## FUNDING SUPPORT

This work was supported by University Hospital, Newark NJ; and the National Institute of Allergy and Infectious Diseases of the National Institutes of Health [grant number T32AI125185].

## Notes

### Competing Interest Statement

The authors have declared no competing interest.

